# Ocular Biometry Profile of Adult Cataract Surgery Candidates at University of Gondar, Tertiary Eye Care and Training Center, North West Ethiopia

**DOI:** 10.1101/2022.07.20.22277853

**Authors:** Bemnet Feleke, Wossen Mulugeta, Teshager Wondale, Asamere Tsegaw

**Author notes:** <u>Correspondence</u> Dr. Asamere Tsegaw.

## Abstract

**Background:** Precise measurement of the radius of curvature [K (mm)] and axial length (AL) of the eye are vital for a good outcome of a cataract extraction surgery. Average values of these dimensions of the optical components depend on race, age and gender. Although there are a number of studies that describe these mean parameters in the Caucasian, Hispanic and Asian populations, there has been little attention given to the African population.

**Objectives:** The aim of this study was to estimate the biometric parameters of patients who had cataract surgery at University of Gondar (UoG), Tertiary eye care and training centre.

**Materials and methods:** Hospital based cross sectional study was done on patients who visited UoG Tertiary Eye Care and Training Center for cataract extraction surgery. Righton retinomax K plus 3 autorefractor,(Hutama Medical, Indonesia) and Pac Scan 300A contact A scan ultrasound, (Sonomed Escalon, NY, USA) were used to measure K (mm) and AL and calculate intraocular lens (IOL) power, respectively. Data were collected using a structured questionnaire, then entered to SPSS version 25 statistical software and analyzed.

**Result:** A total of 838 eyes (433 right eyes and 405 left eyes) of 486 of patients were included in this study. The mean age was 62.5 ±12.88 years. The mean K1 and K2 were 7.75±0.38 mm and 7.48±0.33 mm, respectively. The mean average of K1 and K2 (AVK) was 7.61±0.33 mm. The mean AL measured was 23.06±1.05 mm and the mean Intraocular Lens (IOL) power calculated was 21.12±2.95 D. Significant difference in mean AVK and mean AL of the eye exists between male and female patients, p value =0.000.

**Conclusion:** This study provides the mean values of radius of curvature of cornea, axial length of the eye and IOL power of patients who were about to undergo cataract extraction surgery at the study center. Male patients had steeper corneas and longer axial lengths than females. But age had a variable effect on both AVK and AL.

## Introduction

Ocular biometry refers to the dimensions of the eye. The average values of the dimensions of the optical components depend on race, age and gender. Axial length (AL) of the eye is defined as the distance between the anterior corneal surface and internal limiting membrane of the retina. (1)

In cataract surgery, accurate measurement of axial length (AL) and corneal power or corneal radius of curvature (K), are crucial to achieve satisfactory postoperative refractive results and reduce spectacle dependence.

A retrospective hospital based study done in California, USA in 1993 on 600 eyes showed that the mean axial length was 23.65 (±1.28)mm. Eyes in patients younger than age 65 years (158 eyes) had mean axial length of 24.08 (±1.53)mm, eyes in patients aged from 65 to 75 years (252 eyes) had mean axial length of 23.67 ±1.19 mm, and eyes in patients older than age 75 years (190 eyes) had mean axial length of 23.26 ±1.03 mm. The differences were statistically significant (P<.0001). AL was measured ultrasonically with immersion technique. (2)

A retrospective study of 13,012 eyes of 6,506 patients, who underwent cataract surgery, in the year 2017, was performed at a hospital in Portugal and reported mean AL and K values of 43.91 ± 1.71 D, and 3.25 ± 0.44 mm, respectively. Male patients had longer AL (p < .001) and flatter corneas (p < .001). (3)

A cross-sectional hospital-based study in Southern China included 6750 eyes of 4561 consecutive cataract candidates, between 2007 and 2011. The means of AL and K were 24.07 ± 2.14 mm and 44.13 ± 1.63 D respectively. These values were statistically significantly greater in men than in women (P < .001) and significantly lower with increasing age (P < .001). (4)

In Korea a retrospective study was done that included 800 eyes of 800 patients who underwent cataract surgery from March 2013 to May 2014. The average AL and K value were 23.59 ± 0.89 mm, and 44.16 ± 1.40 D, respectively. Increasing age had a significant association with shorter AL (P < 0.001), but the association between age and K was not significant (P = 0.398).(5)

A prospective cross-sectional study carried out in Nigeria had a total of 130 participants with 77 males and 53 females. Females had slightly higher mean average corneal curvature (AVK) than males, with males having a mean ±SD of 42.83 ± 1.19 Diopter (D) and range of 39.38D-45.38D and females having mean± SD of 43.19 ± 1.17D and range of 40.63D-45.75D. However, age and gender did not significantly affect horizontal, vertical or average corneal curvature.(6)

A cross-sectional study undertaken between June 2018 and July 2019 at St. Paul’s Hospital, Ethiopia included 400 eyes of 200 subjects. The mean ±SD of average corneal refractive power and axial length (AL) were 43.94 ± 9.78 diopter and 22.96 ± 0.82 mm, respectively. (7)

Each year about 4000 cataract surgeries are done at UOG Tertiary eye care and training center. For each of these patients, corneal radius of curvature, axial length and IOL power is calculated before cataract surgery. But for patients for whom such measurement cannot be done due to various reasons an average IOL power calculated for western population is inserted after cataract surgery. This is because the mean values of the biometric profile of our population is not known, which can lead to unexpected refractive result in the postoperative period. This study tried to determine the means of K, AL and IOL power of cataract surgery candidate patients at the center. It has also tried to describe the relationship between age and sex with these variables.

## Patients and Methods

### Study design and period

Institution based Prospective cross-sectional study was carried out from September 2019-August 2020 at UoG, Tertiary eye care and training center, NW Ethiopia.

### Study area

The study was conducted at UoG Tertiary Eye Care and Training Center, a major eye care and training center in Ethiopia. It is an ophthalmic referral center for an estimated 14 million people living in North-West Ethiopia. The center provides eye care services both at base hospital and rural outreach sites. The base hospital has 8 out-patient clinics, facilities for in-patient care with 30 beds and five operation theatres.

#### Study population

All adult patients above the age of 16 who were candidates for cataract extraction surgery and for whom biometry was measured at university of Gondar tertiary eye care and training center were study patients.

#### Exclusion criteria

A pseudophakic eye

Eyes with traumatic cataract

corneal disease or ocular surface disorders that can affect corneal curvature, such as, pterygium, corneal degeneration or dystrophy

history of ocular inflammation

### Operational definitions

**Axial length-** the distance between the anterior corneal surface and internal limiting membrane of the retina.In emmetropic eyes axial length measurements ranging from 22 to 25 mm are considered normal.

**Corneal radius of curvature –** Keratometric measurements of the primary meridians average 7.8 mm, with a large range from 7.0 mm to 9.5 mm for normal corneas. A conversion to keratometric diopters can be quickly performed by dividing 337.5 by the radius of curvature in millimeters, and, likewise, a conversion to radius of curvature (in mm) can be done by dividing 337.5 by the surface power in keratometric diopters.

### Data Collection Procedures

At the outpatient clinics ophthalmic history, review of systems, visual acuity, intraocular pressure, detailed slit lamp examination of the anterior segment and when possible posterior segment examination were done for each cataract surgery candidate patient by senior ophthalmologists. Biometry was done by senior residents.

Data were collected using a standardized checklist and data extraction format which consisted of demographic data, presenting visual acuity and Biometry measurements.

#### Visual Acuity

Tumbling-E Snellen’s charts were used to assess visual acuity, starting the examination with the patient at 6m distance from the chart in a well illuminated room. Grading of visual impairment and blindness was done according to the WHO classification system as follows: visual acuity better or equal to 6/18 – normal; visual acuity less than or equal to 6/24 and better than 6/60 – moderate visual impairment; visual acuity less than or equal to 6/60 and better than counting fingers at 3m – severe visual impairment; visual acuity less than or equal to counting fingers at 3m – blindness;.

### Biometry Measurement techniques

Corneal radius of curvature was measured using the Rightonretinomax K plus 3 autorefractor, Hutama Medical, Indonesia. During the measurement patients were instructed to look straight into the instrument’s screen at its center while the technician looks through the eyepiece. The aim was for the technician to center the square at the center of the dots when the mires are well focused.

When the circles of dots (mires) are well focused and the alignment is correct, the instrument takes measurement of the radius of curvature in the steep meridian (K1 mm) and the flat meridian (K2 mm). Similar procedure was repeated for the other eye. The result of both eyes will appear on the screen through the eyepiece, which was then copied to the data collection sheet.

The next step was to insert patient data and K1 (mm) and K2 (mm) numbers into the Pac Scan 300A,, Sonomed Escalon, NY, USA, in order to measure the AL and IOL power. With the patient comfortably seated, topical anesthetic drops were instilled into the eyes. Then keeping the A scan probe as perpendicular to the corneal surface as possible and trying not to put pressure on the cornea, five measurements were taken with the maximum tolerable deviation being 0.06. Measurement was repeated if there was >0.3mm AL and 2D in IOL power difference between the two eyes.

The A-constant was set to 118.5 and the IOL power was calculated automatically by the instrument and displayed on the screen. The AL in millimeters (mm) and IOL power in diopter (D) were then recorded.

### Data processing and analysis

The collected data were checked for accuracy, cleaned, coded and entered into Statistical Package for Social Sciences (SPSS) version 24. Descriptive analyses were computed to calculate mean and standard deviation of AL, corneal radius of curvature and IOL power. Variation of the means of these parameters according to age and sex were evaluated using one-way analysis of variance (ANOVA) and t-test. Results were presented in the form of tables and figures.

### Ethical considerations

The study was conducted in full compliance with the 1964 Helsinki declaration on research involving human subjects. Prior to commencement of the study, ethical clearance was obtained from the Ethics Committee (Institutional Review Board) of University of Gondar and Informed consent was obtained from each study subject.

## Results

### Socio-demographic Characteristics

A total 838 eyes (433 right eyes and 405 left eyes) of 486 study participants were included in this study. The mean age of the study participants was 62.5+/-12.88 years (range 17-89 years). Thirty-one-point nine percent of the study participants were age group between 61-70 years and 57.6% of the study participants were male. Almost three-fourth, 362 (74.5 %) of the participants live in rural areas and 57.2% were farmers. (Table-I)

**Table 1.**
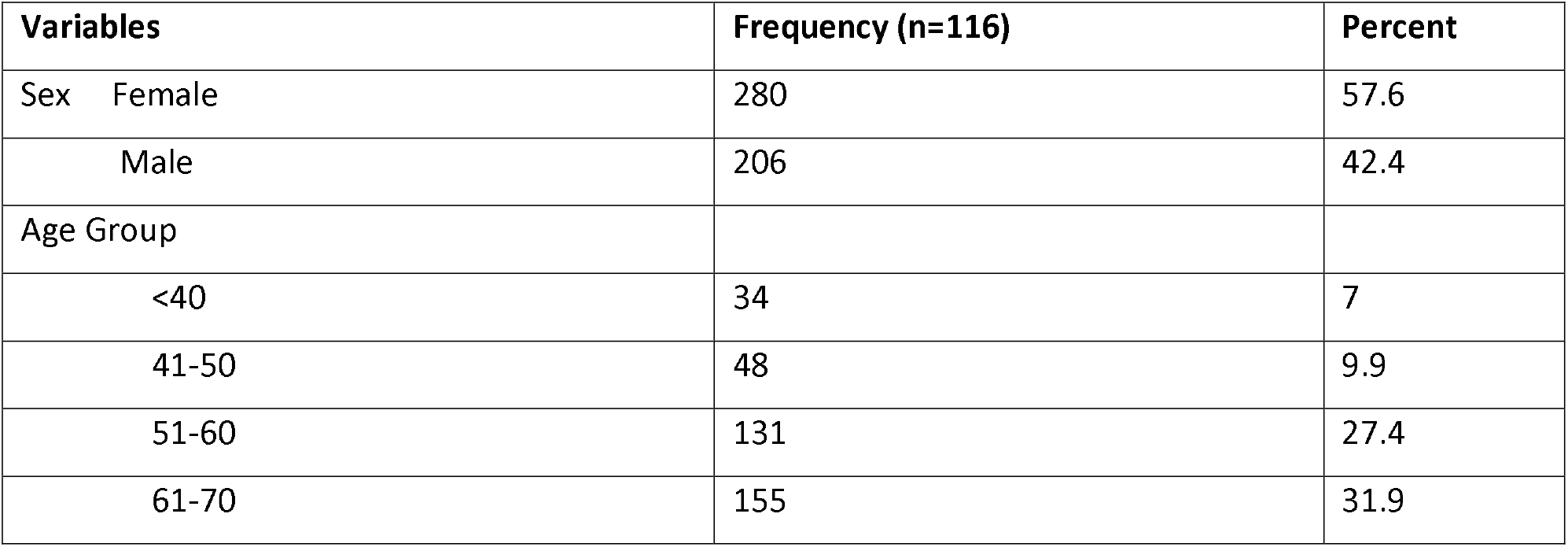
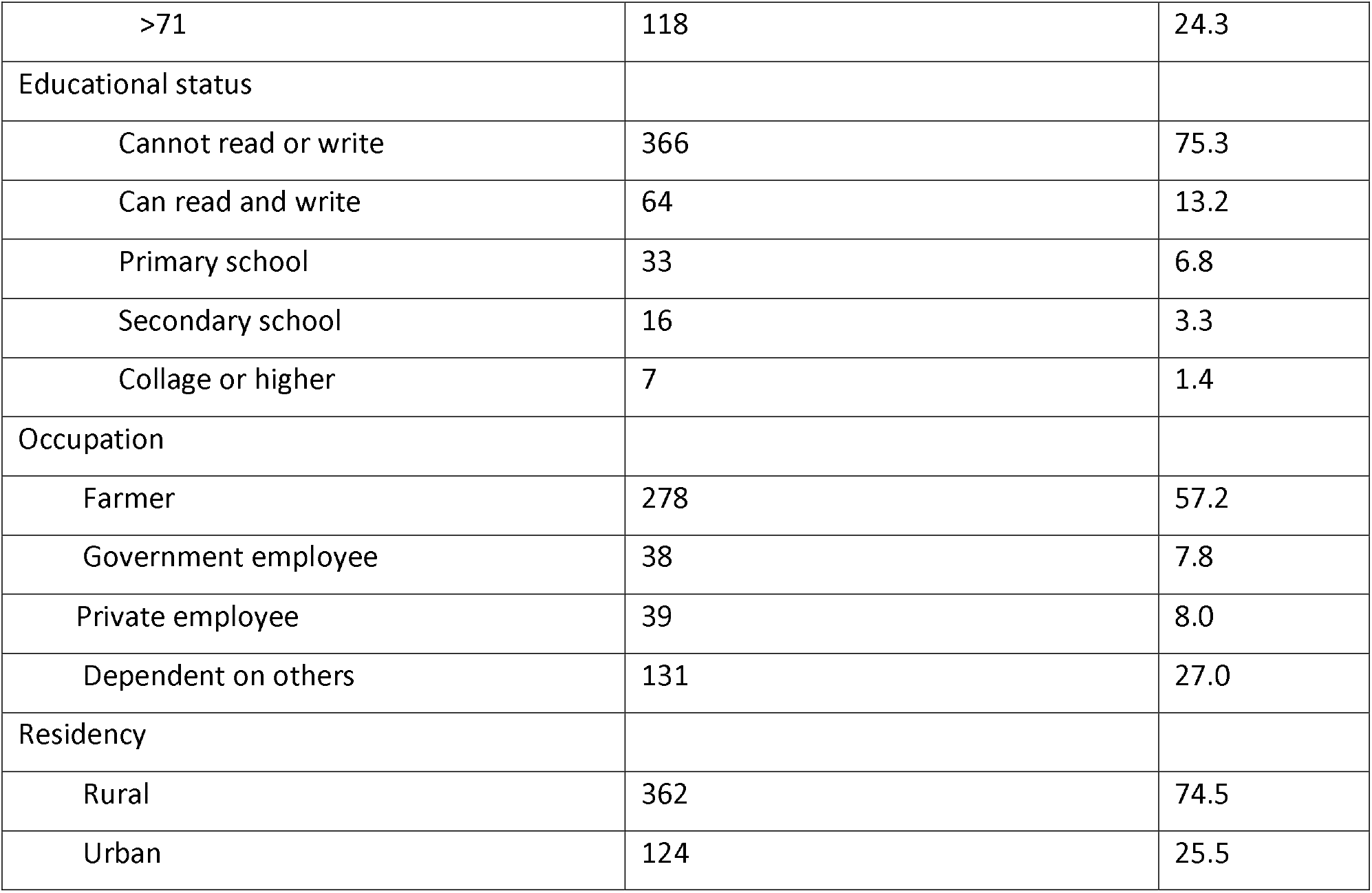
Socio-demographic characteristic of cataract surgery candidate patients who visited UOGTECTC in Gondar town Northwest Ethiopia, 2020 (n=486)

| Variables | Frequency (n=116) | Percent |
| --- | --- | --- |
| Sex Female | 280 | 57.6 |
| Male | 206 | 42.4 |
| Age Group |  |  |
| <40 | 34 | 7 |
| 41-50 | 48 | 9.9 |
| 51-60 | 131 | 27.4 |
| 61-70 | 155 | 31.9 |
| >71 | 118 | 24.3 |
| Educational status |  |  |
| Cannot read or write | 366 | 75.3 |
| Can read and write | 64 | 13.2 |
| Primary school | 33 | 6.8 |
| Secondary school | 16 | 3.3 |
| Collage or higher | 7 | 1.4 |
| Occupation |  |  |
| Farmer | 278 | 57.2 |
| Government employee | 38 | 7.8 |
| Private employee | 39 | 8.0 |
| Dependent on others | 131 | 27.0 |
| Residency |  |  |
| Rural | 362 | 74.5 |
| Urban | 124 | 25.5 |

### Clinical Characteristics

The visual acuity measurement was hand motion or light perception in 43% (359) of studied eyes: 39.3% (191) in the right and 34.6% (168) in the left eyes. (Figure 1)

**Figure 1.**
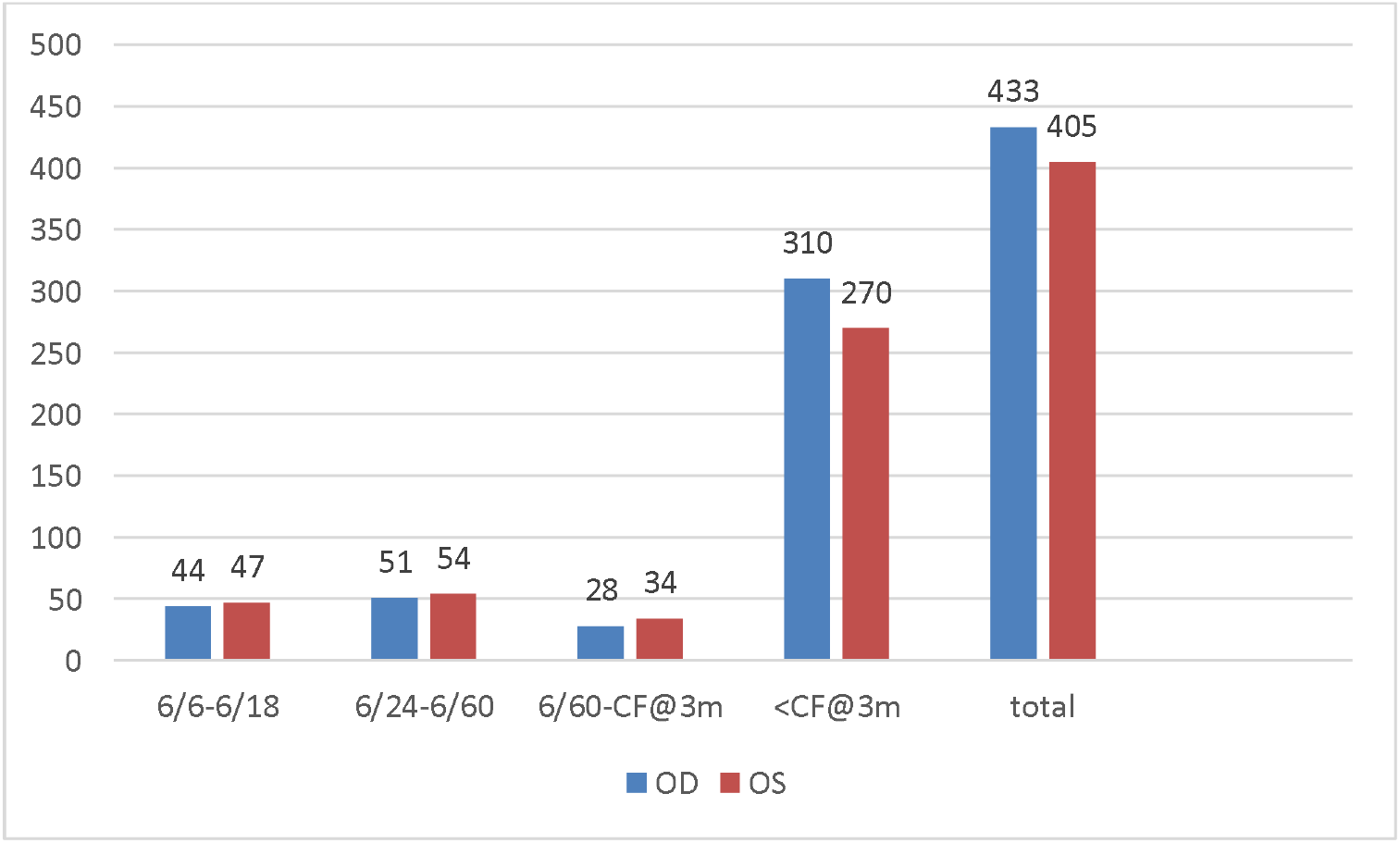
Presenting visual acuity of right eyes and left eyes of cataract surgery candidate patients who visited UOGTECTC in Gondar town. Northwest Ethiopia, 2020

The mean vertical and horizontal corneal curvatures (K1 and K2) were 7.75±0.385 mm (43.65 ±2.14 D) and 7.48±0.339 mm (45.2 ±1.99 D), respectively. The mean average corneal curvature (AVK) was 7.61±0.337mm (44.42 ±1.92 D). The mean axial length measured was 23.06±1.059 mm and the mean IOL power calculated was 21.12 ±2 .95 D. (Table-2)

**Table 2.** Mean, standard deviation (SD) and range of measured parameters of cataract surgery candidate patients who visited UOGTECTC in Gondar town Northwest Ethiopia, 2020

| Variable | Mean | Standard Deviation | Min-Max |
| --- | --- | --- | --- |
| Vertical Corneal Curvature | 7.75mm | 0.385 | 6.74 - 10.74 mm |
|  | 43.65 D | 2.14 | 32.45-50.0 D |
| Horizontal Corneal Curvature | 7.48mm | 0.339 | 6.25 - 8.94 mm |
|  | 45.2 D | 1.99 | 37.75- 54.0 D |
| Average Corneal Curvature | 7.61mm | 0.337 | 6.55 - 9.07 |
|  | 44.42 D | 1.92 | 37.21- 51.52 D |
| Axial Length | 23.06mm | 1.059 | 18.75 - 30.15mm |
| IOL Power | 21.09 D | 0.006 | -6 - 29.5 D |

While comparing the means of the ocular biometry values between male and female, male participants had longer corneal radii of curvature, longer axial lengths and had a calculated value of IOL power that is lesser than female participants. (Table-3)

**Table 3.** Comparison of means of measured parameters based on gender of cataract surgery candidate patients who visited UOGTECTC in Gondar town. Northwest Ethiopia, 2020

| Outcome variables | Mean± SD |  |
| --- | --- | --- |
|  | Male | Female |
| Vertical Corneal Curvature | 7.8084 ± 0.3935 mm | 7.6769 ± 0.3612 mm |
|  | 43.37 ± 2.17 D | 44.06 ± 2.04 D |
| Horizontal Corneal Curvature | 7.5352 ± 0.3233 mm | 7.3954 ± 0.3419 mm |
|  | 44.89 ± 1.9 D | 45.76 ± 2.1 D |
| Average Corneal Curvature | 7.6708 ± 0.3304 mm | 7.5304 ± 0.3281 mm |
|  | 44.08 ± 1.9 D | 44.89 ± 1.9 D |
| Axial Length | 23.2136 ± 0.9197 mm | 22.8640 ± 1.1940 mm |
| IOL Power | 20.9841 ± 2.5751 D | 21.2968 ± 3.3778 D |

Across all age groups AVK was within the range of 7.51-8.00mm (42.18-44.94D), except in patients of age 41-50 who had an AVK within 7.01-7.50mm (45- 48.4 D) range in 46.9% of them. This means patients in later age group had slightly steeper corneas than other age groups. (Table-4) (Table 5)

**Table 4.**
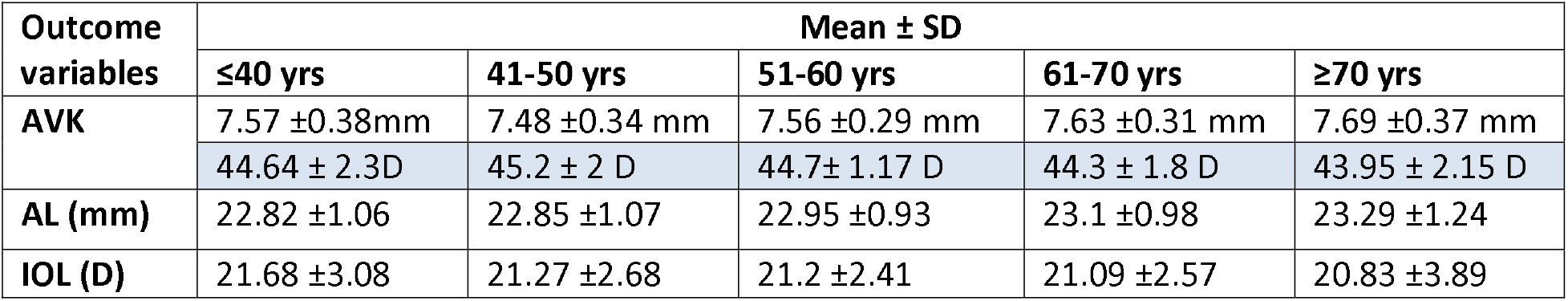
Comparison of means of measured parameters based on age category of cataract surgery candidate patients who visited UOGTECTC in Gondar town. Northwest Ethiopia, 2020

| Outcome variables | Mean ± SD |  |  |  |  |
| --- | --- | --- | --- | --- | --- |
|  | ≤40 yrs | 41-50 yrs | 51-60 yrs | 61-70 yrs | ≥70 yrs |
| AVK | 7.57 ±0.38mm | 7.48 ±0.34 mm | 7.56 ±0.29 mm | 7.63 ±0.31 mm | 7.69 ±0.37 mm |
|  | 44.64 ± 2.3D | 45.2 ± 2 D | 44.7± 1.17 D | 44.3 ± 1.8 D | 43.95 ± 2.15 D |
| AL (mm) | 22.82 ±1.06 | 22.85 ±1.07 | 22.95 ±0.93 | 23.1 ±0.98 | 23.29 ±1.24 |
| IOL (D) | 21.68 ±3.08 | 21.27 ±2.68 | 21.2 ±2.41 | 21.09 ±2.57 | 20.83 ±3.89 |

**Table 5.** Frequency and percentage of AVK by gender of the cataract surgery candidate patients who visited UOGTECTC in Gondar town, Northwest Ethiopia, 2020

|  |  | Sex |  |
| --- | --- | --- | --- |
|  |  | Male N(%) | Female N(%) |
| AVK | 6.51-7.00mm (48.21- 51.84D) | 8(32) | 17(68) |
|  | 7.01-7.50mm (45- 48.14 D) | 134(45.9) | 158 (54.1) |
|  | 7.51-8.00mm (42.18- 44.94D) | 276(63.9) | 156(36.1) |
|  | ≥8.01mm (≥ 42.13D) | 61(68.5) | 28(31.5) |

A majority of male patients 63.9% (n=276) had AVK in the range of 7.51-8.00mm (42.18- 44.94D). Almost equal number of female patients had AVK within the range of 7.01-7.50mm (45- 48.4 D) and 7.51-8.00mm (42.18- 44.94D). (Table 6)

**Table 6.** Frequency and percentage of AL by age groups of cataract surgery candidate patients who visited UOGTECTC in Gondar town, Northwest Ethiopia, 2020

| Axial length |  | Age (years) |  |  |  |  |
| --- | --- | --- | --- | --- | --- | --- |
|  |  | ≤40 N (%) | 41-50 N (%) | 51-60 N (%) | 61-70 N (%) | ≥71 N (%) |
|  | 18.01-20 mm | 0(0.0) | 0(0.0) | 0(0.0) | 0(0.0) | 1(0.5) |
|  | 20.01-22 mm | 14(24.6) | 16(19.8) | 27(11.7) | 27(10.0) | 12(6.0) |
|  | 22.01-24 mm | 37(64.9) | 53(65.4) | 183(79.6) | 209(77.7) | 155(77.1) |
|  | ≥24.01 mm | 6(10.5) | 12(14.8) | 20(8.7) | 33(12.3) | 33(16.4) |

A majority of male and female patients, 59.8% and 40.2% respectively, have an AL within the range 22.01-24mm and within all age groups the commonest range of AL was 22.01-24.00mm. (Table-7) and (Table 8)

**Table 7.** Frequency and percentage of AL by gender of the cataract surgery candidate patients who visited UOGTECTC in Gondar town, Northwest Ethiopia, 2020

|  |  | sex |  |
| --- | --- | --- | --- |
|  |  | Male N(%) | Female N(%) |
| AL | 18.01-20 mm | 1 | 0 |
|  | 20.01-22 mm | 30(31.3) | 66(68.8) |
|  | 22.01-24 mm | 381(59.8) | 256(40.2) |
|  | ≥24.01 mm | 67(64.4) | 37(35.6) |

**Table 8.** Frequency and Percentage of IOL power by age group of the cataract surgery candidate patients who visited UOGTECTC in Gondar town. Northwest Ethiopia, 2020

| Intraocular Lens |  |  | Age |  |  |  |  | total |
| --- | --- | --- | --- | --- | --- | --- | --- | --- |
|  |  |  | ≤40<br>N (%) | 41-50<br>N (%) | 51-60<br>N (%) | 61-70<br>N (%) | ≥71<br>N (%) |  |
|  | ≤18 |  | 2(3.5) | 6(7.4) | 22(9.6) | 13(4.8) | 23(11.4) | 66 (7.9) |
|  | 18.01-20.00 D |  | 12(21.1) | 15(18.5) | 32(13.9) | 64(23.8) | 38(18.9) | 161 (19.2) |
|  | 20.01-22.00 D |  | 21(36.8) | 33(40.7) | 107(46.5) | 116(43.1) | 80(39.8) | 357 (42.6) |
|  | 22.01-24.00 D |  | 12(21.1) | 20(24.7) | 52(22.6) | 61(22.7) | 46(22.9) | 191 (22.8) |
|  | ≥24.01 D |  | 10(17.5) | 7(8.6) | 17(7.4) | 15(5.6) | 14(7) | 63 (7.5) |

Out of 838 eyes 42.6% (n=357) were calculated to have an IOL power within the range 20.01-22.00D. One hundred ninety one (22.8%) and 161 (19.2%) eyes were calculated to have an IOL power within the ranges 22.01-24.00D and 18.01-20.0D, respectively.(Table 9)

**Table 9.** Independent sample t-test for means of measured ocular biometry parameters based on the gender of the cataract surgery candidate patients who visited UOGTECTC in Gondar town. Northwest Ethiopia, 2020

| Outcome variables | t value | p value | Mean difference | 95%CI for MD |  |
| --- | --- | --- | --- | --- | --- |
|  |  |  |  | Lower | Upper |
| AVK | 6.103 | 0.000 | 0.14035 | 0.09521 | 0.18549 |
| AL | 4.788 | 0.000 | 0.34961 | 0.20627 | 0.49294 |
| IOL Power | -1.52 | 0.129 | -0.31266 | -0.71629 | 0.09096 |

Independent sample t-test revealed that a significant difference in mean AVK and mean AL of the eyes exists between male and female patients: t values of both AVK and AL with gender were statically significant with p value =0.000. The mean IOL power of the eyes in males and females have no statistically significant difference (with p value= 0.129). (Table 10)

**Table 10.** ANOVA test between measured ocular biometry parameters and age groups of the cataract surgery candidate patients who visited UOGTECTC in Gondar town. Northwest Ethiopia, 2020

| Outcome Variables |  | Sum of Squares | Df | Mean Square | F | Sig. |
| --- | --- | --- | --- | --- | --- | --- |
| Average corneal curvature | between Groups | 3.455 | 4 | 0.864 | 7.878 | 0.000 |
|  | Within Groups | 91.32 | 833 | 0.11 |  |  |
|  | Total | 94.775 | 837 |  |  |  |
| Axial length | Between Groups | 21.127 | 4 | 5.282 | 4.789 | 0.001 |
|  | Within Groups | 918.765 | 833 | 1.103 |  |  |
|  | Total | 939.892 | 837 |  |  |  |
| Intra ocular lens power | Between Groups | 38.079 | 4 | 9.520 | 1.096 | 0.357 |
|  | Within Groups | 7236.319 | 833 | 8.867 |  |  |
|  | Total | 7274.399 | 837 |  |  |  |

Before we had handled ANOVA, assumptions of ANOVA were checked and all the assumptions were satisfied. The data was checked for assumptions priory. Normality check performed with the Shapiro-Wilk test revealed that the data was normally distributed.

The result indicated that there was a significant difference in the means AVK and AL of the eyes between different age groups F(4, 833)=7.878, p<0.001 and F(4, 833)=4.789, p<0.002, respectively. But there was no significant difference in mean IOL power between the different age groups F (4, 833)= 1.096, p=0.357.

Post-hoc analysis was conducted to compare means of AVKs of specific age groups. The result of the post-hoc analysis with the Bonferroni test indicated that there was a significant difference between age group 41-50 VS 61-70, 41-50 VS ≥71 and age group 51-60 Vs >71 with p-value of 0.005, 0.000 and 0.000 respectively. After the age of 50 years, the cornea shows flattening with increasing age. (Table 11)

Similar procedures were followed to compare means of ALs of the eyes of the specific age groups. The result indicated that there was a significant difference between age group ≤40 VS ≥71, 41-50 VS ≥71 and 51-60 VS ≥71 with p-value of 0.021, 0.011 and 0.007 respectively. This leads us to the conclusion, with increasing age, AL of eyes also increased. (Table 11)

## Discussion

The mean age of the study participants was 62.5 +/- 12.88 years with range between 17 and 89. This result is comparable to the result found in Korea where the mean age was 67.4 ±12.0 years with range between 20–94 years (5). Studies done on similar population or patients who were about to undergo cataract extraction surgery in Central and Southern china had older patients, 70.51 ± 9.81 range 32-95 years and 70.4 ± 10.5 years (range 40-101 years), respectively (4,8). This mean age difference may be due to the difference in life expectancy between Asian population and ours.

The study done in Addis Ababa Ethiopia had a mean age of 40.31± 11.39 with a range from 18 to 69. The reason that their mean age is less than ours is that they excluded patients who had age related cataract that made fundus examination impossible (7).

The mean average corneal curvature (AVK) of our study population was found to be 7.61± 0.33mm, which is almost equal to the result found in Singapore, 7.61 ± 0.26 mm (9). Similar results were also found from Southern China and Portugal, which were 7.65 ± 0.16 mm (4) and 7.69 ± 0.3mm (3), respectively. A study from Nigeria had a longer AVK measurement, 7.81 ± 0.28mm (10). An even longer AVK, 8.07± 1.79mm, was reported by a study done in Addis Ababa, Ethiopia (7).

In our study, AVK in males was 7.67 ± 0.33 mm and in females 7.53 ± 0.32 mm. Studies from Portugal, Southern China Nigeria and Addis Ababa had slightly larger values than ours, 7.76 ± 0.19 mm in men and 7.63±0.22 mm in females in Portugal (3), 7.71± 0.28mm in men and 7.61± 0.27 mm in females in Southern China(4) and 7.88 ± 0.22 mm in male and 7.81± 0.21 mm in female in Nigeria (10) and 7.93± 0.23mm in males and 7.62± 0.53mm in females in Addis Ababa (7).

When comparing AVK between the age groups, this study showed that age group less than 40 years had a longer AVK than age group between 40 and 50 years, then the mean continued to increase as age increases. In the study done in Singapore, the mean AVK between the age groups was within close range and the difference was not statistically significant, p=0.22 (9). The study from South China had a different result. It had a decreasing pattern from the age group 40-49 years to the age group 70-79 years then it showed an increment (4).

The Mean axial length of our participants was 23.06 ±1.05mm, which is similar to the result found in Addis Ababa 22.96 ±0.82mm (7) Iran 23.14 mm (15), in Nigeria 23.78 ±0.91mm (10) and in Portugal 23.87± 1.55mm (3). The result from South China was longer than our result, 24.07 ± 2.14 mm(4). In the above studies and also in a study from Singapore (9), males had significantly longer axial lengths than female, as it is in ours. But in a study done in Korea the ALs between male and female were almost equal, 23.59 ± 0.90 mm in men and 23.60 ±0.89 mm in women (5).

In our study the mean AL increased with increasing age. However, studies from Iran (11), Singapore (9) and South China (4) had opposite results. In these studies AL decreased with increasing age. A study from South Africa showed that AL increased from the age group 10-19 years to the age group 30-39 years then decreased until the group 50-59 years and increased after the age group above 60 years (12).

## Conclusion

This study provides the mean values of radius of corneal curvature, axial length and IOL power of patients who were about to undergo cataract extraction surgery at UoG Tertiary eye care and training center. Male patients had steeper corneas and longer axial lengths than females. But age had a variable effect on both AVK and axial length.

## Data Availability

All relevant data are
within the manuscript and its Supporting
Information files.

---

**Table 4.** Frequency and percentage of AVK by age groups of cataract surgery candidate patients who visited UOGTECTC in Gondar town. Northwest Ethiopia, 2020

| Average corneal | Age |  |  |  |  |
| --- | --- | --- | --- | --- | --- |
|  | ≤40 N | 41-50 N | 51-60 N | 61-70 N | ≥71 N (%) |

| curvature (AVK) |  | (%) | (%) | (%) | (%) |  |
| --- | --- | --- | --- | --- | --- | --- |
|  | 6.51-7.00 mm<br>(48.21- 51.84D) | 7(12.3) | 5(6.2) | 5(2.2) | 4(1.5) | 4(2.0) |
|  | 7.01-7.50 mm<br>(45- 48.14 D) | 18(31.6) | 38(46.9) | 90(39.1) | 87(32.3) | 59(29.4) |
|  | 7.51-8.00 mm<br>(42.18- 44.94D) | 23(40.4) | 32(39.5) | 124(53.9) | 151(56.1) | 103(51.2) |
|  | ≥8.01 mm<br>(≥ 42.13D) | 9(15.8) | 6(7.4) | 11(4.8) | 27(10.0) | 35(17.4) |

## Notes

### Competing Interest Statement

The authors have declared no competing interest.

### Funding Statement

No funding for this study

